# Changes in the medical admissions and mortality amongst children in four South African hospitals following the COVID-19 pandemic : A five-year review

**DOI:** 10.1101/2023.12.28.23300622

**Authors:** Naidoo Kimesh Loganathan, Dorward Jienchi, Chinniah Kogielambal, Lawler Melissa, Nattar Yugendhree, Bottomley Christian, Archary Moherndran

## Abstract

**Background:** Vulnerable children from poor communities with high HIV and Tuberculosis(TB) burdens were impacted by COVID-19 lockdowns. Concern was raised about the extent of this impact and anticipated post-pandemic surges in mortality.

**Methods:** Interrupted time series segmented regression analyses were done using routinely collected facility-level data of children admitted for medical conditions at four South African referral hospitals. Monthly admission and mortality data over a 60-month period from 01 April 2018 to 31 January 2023 was analysed using models which included dummy lockdown level variables, a dummy post-COVID period variable, Fourier terms to account for seasonality, and excess mortality as a proxy for healthcare burden.

**Results:** Of the 45 015 admissions analysed, 1237(2·75%) demised with significant decreases in admissions during all the lockdown levels, with the most significant mean monthly decrease of 450(95%, CI=657·3, −244·3) p<0·001 in level 5 (the most severe) lockdown. There was evidence of loss of seasonality on a six-month scale during the COVID periods for all admissions (p=0·002), including under-one-year-olds (p=0·034) and under-five-year-olds (p=0·004). No decreases in mortality accompanied decreased admissions. Post-pandemic surges in admissions or mortality were not identified in children with acute gastroenteritis, acute pneumonia and severe acute malnutrition.

**Conclusion:** During the COVID-19 pandemic, paediatric admissions in 4 hospitals serving communities with high levels of HIV, TB and poverty decreased similar to global experiences however there was no change in in-hospital mortality. No post-pandemic surge in admissions or mortality were documented. Differences in the impact of pandemic control measures on transmission of childhood infections and access to health care may account for differing outcomes seen in our setting compared to the global experiences. Further studies are needed to understand the impact of pandemic control measures on healthcare provision and transmission dynamics and to better inform future responses amongst vulnerable child populations.

## Background

The national lockdown regulations promulgated across the globe due to the COVID-19 pandemic disrupted essential healthcare services for most populations [1]. Healthcare visits, including emergency outpatient visits and admissions, decreased sharply among children in all countries, especially between February 2020 and December 2021[2–5]. Decreases of 19%, 50% and 56% in paediatric admissions were documented in Cameroon, South Africa (SA) and across Europe, respectively, compared with pre-COVID-19 time periods [3–5]. Vulnerable populations, including children in lower- and middle-income countries (LMICs), were especially negatively impacted due to these lockdown measures [2–4].

The decrease in paediatric admissions documented has been more marked in children with communicable (77%) compared with non-communicable diseases (37%) [5].Children with lower respiratory tract infections (LRTI), which includes those with highly communicable viral bronchiolitis, also decreased [6–8]. Changes in seasonal patterns of viral bronchiolitis when compared with patterns in identified in previous pre-COVID-19 years were noted [6]. This was postulated to occur due to reduced person-to-person transmission, and it raised concerns that a rebound would occur when transmission mitigating strategies and lockdown levels were curtailed [6].

Visits to children’s preventative health services decreased significantly across multiple countries, especially after the start of the COVID-19 pandemic in February 2020 [9]. These decreases were documented in both urban and rural primary healthcare facilities.^10^ Outpatient visits for children with Human Immunodeficiency Virus (HIV) dropped by 41%, and antiretroviral treatment initiation of newly diagnosed children also decreased in 2020 and 2021[11]. These changes in access and utilisation of both preventative healthcare and HIV chronic care raised concerns for negative health consequences, especially where poverty, HIV and Tuberculosis (TB) is common and where many live in high density communities [11]. HIV viral suppression rates, however, were shown to be maintained among children, suggesting some chronic disease programmes remained reasonably robust over this period [12].

Overall, the COVID-19 pandemic disrupted healthcare provision and health-seeking behaviour and was postulated to disproportionately impact specific subpopulations in low-income countries with fragile health systems and pervasive social-structural vulnerabilities[13]. Documentation of these indirect effects of the COVID-19 pandemic has been largely restricted to the period during the peaks of the COVID-19 lockdowns between February 2020 and December 2021 and not adequately documented in communities with high burdens of HIV, Tuberculosis (TB) and malnutrition [11]. With this disruption of healthcare, concern was raised about the deleterious effects on mortality and morbidity rates rising, specifically in these vulnerable children in such communities after the removal of lockdown measures [14].

Children hospitalised in specialist referral hospitals include complex cases requiring higher levels of medical care and support [15]. This study describes and analyses changes in admission and in-hospital mortality amongst children in South African specialist referral hospitals during all the levels of national lockdowns associated with the COVID-19 pandemic and the post-pandemic period.

## METHODS

### Study design and population

We conducted an interrupted time series analysis of routinely collected facility-level data of children below the age of 13 years hospitalised across all four of the largest public sector ( non-fee-paying) specialist referral hospitals in the city of Durban (eThekwini District), Kwa Zulu-Natal(KZN). The data collected included those hospitalised with medical diagnoses only, thus allowing analysis to reflect on the impact of the COVID-19 pandemic, specifically on communicable diseases. In-born neonates and children hospitalised for surgical ( general surgery, trauma, ear nose and throat procedures, orthopaedic reasons) or other non-medical reasons (psychiatric and social admissions for respite care or neglect) were purposefully excluded from the analysis.

We used data from the King Edward VIII, Mahatma Gandhi Memorial, Prince Mshyeni Memorial and R K Khan Memorial hospitals, which provide 240 in-patient paediatric medical specialist care beds (including designated high care and beds for interim invasive ventilation) for approximately 1,1 million children [15,16]. The children admitted to these hospitals are referred by primary healthcare providers (nurse-run day-clinics, family practitioners, non-specialist district hospitals) and are generally complex cases requiring higher care levels. Children who require longer-term invasive ventilation (>72 hours) are referred to paediatric intensive care units located at the quaternary hospital. The majority of the children who attend and are hospitalised in these four referral hospitals are from lower socio-economic communities and live in communities with high population densities [15]. The HIV antenatal seroprevalence of the population served by these hospitals is high at 44·3%(CI;41·6-46·7), reflecting a high burden of both HIV-exposed infants and HIV-infected children [16,17].

The study period spanned 60 months and included 23 months in the pre-COVID-19 period (01 April 2018 to 28 February 2020), 23 months of the designated COVID-19 period (01 March 2020 to 31 January 2022), during which one of the five lockdown stages were promulgated and 14 months post COVID-19 period (01 February 2022 to 31 January 2023), when no lockdowns were in place. South Africa announced a national lockdown on 23 March 2020, implemented on 27 March 2020 [18,19]. Starting at level 5, the lockdown was one of the most severe globally, with restrictions on movement and cancellation of public transport, although travel to receive healthcare was allowed [19]. The lockdown was eased to level 4 on May 1, 2020, when public transport was allowed, and to level 3 on 01 June 2020, which allowed some economic activity to resume. Over the 23-month COVID-19 period, lockdown levels vacillated depending on the authorities’ anticipated need to prevent community transmission [18,19]. Monthly data in the COVID period were thus stratified according to the predominant lockdown level in each of the 23 months in this period.

### Data collection

The admission diagnosis of children included in the facility-level monthly data was obtained from in-patient records that an attending paediatrician validated. Data on hospitalised children included children in all age groups below 13 years of age (SA’s referral hospitals have a 13-year-old cut-off for paediatric care), those below one year of age (infant) and those between one and five years of age. Data on hospitalised children under the age of five years with lower respiratory tract infections (LRTI) or acute gastroenteritis (AGE) as their main diagnosis were specifically tracked. In this study, the term LRTI as a diagnostic category includes patients with lobar or bronchopneumonia, bronchiolitis and bronchitis. This categorisation was based on a standardised nomenclature used by clinicians across all sampled hospitals in admission diagnoses and mortality classification. LRTI excludes upper respiratory tract infections (URTI) or upper airway obstruction, asthma or recurrent wheezing [20]. In addition, monthly admission numbers of children categorised as having severe acute malnutrition (SAM) using the WHO guidelines were also collected. In all four hospitals, the categorisation of a child under five years of age with SAM is verified by a paediatrician and then independently corroborated by an attending dietician within 72 hours post-admission. This dual verification for nutritional categorisation enables weights post-rehydration to be utilised and for lengths or heights to be rechecked for accuracy. In the WHO nutritional classification system, children are classified as either having severe acute malnutrition (SAM), moderate acute malnutrition (MAM), not acutely malnourished but considered at risk (NAM@risk), or not acutely malnourished (NAM) or as overweight or obese [21,22]. The SAM definition was based on weight-for-length z score and/or the presence of nutritional oedema as documented by an attending paediatrician [22]. The mid-upper arm circumference (MUAC) scores were not used in this study as the documentation was inconsistent in the reviewed source documents[21,22]. The numbers of children who demised monthly in all age categories and specifically those with a diagnosis of LRTI, AGE or SAM under the age of five years were also collected.

#### Verification of data

Four independent databases were utilised over the study periods.^23^ These databases corroborated and validated information and ensured minimal missing data. Each hospital’s paediatric department has an in-hospital database used as the primary database. A specialist paediatrician in each hospital is responsible for verifying and entering all weekly admissions tallies and death information (categorised by age and diagnosis) from original case records into this primary database. Admission and mortality data is also verified monthly by paediatricians in the department from a standardised admission and deaths daily register and then submitted to a facility information officer, which feeds this data to a central district-wide district health information system database (DHIS) [23]. In this study, we validated the DHIS data obtained with source data in each hospital from the primary database that the attending paediatricians held to avoid inconsistencies. The third database was the Child Healthcare Problem Identification Programme (Child PIP). Paediatric departments across many SA hospitals utilise this database to record and systematically review child deaths independently, emphasising assessing modifiable factors related to these deaths[24]. Mortality figures per hospital were corroborated using the Child PIP and DHIS and verified at each hospital. The fourth database used verified nutritional categorisation of all in-hospital patients, and in-hospital dietitians maintained these databases in each hospital. The databases were rechecked and then verified with the hospital records for discrepancies.

### Data analysis and interpretation

We used descriptive statistics to summarise data and present summaries of admission, mortality and case fatality rates before, during and after the COVID-19 period with lockdowns. We did an interrupted time series segmented regression analysis by fitting linear regression models with the outcome of monthly paediatric admissions. The models included dummy lockdown level variables indicating 1 or 0 for each level 1 (least severe) to 5 (most severe) of lockdown and a dummy variable for the post-COVID-19 period. COVID-19 waves could also have caused an increased burden on the healthcare system, which may have affected paediatric healthcare use and admissions independently from lockdowns. We, therefore, modelled this by including a continuous variable for excess mortality in eThekwini for each month as a proxy for COVID-19-related burden on the healthcare system. To account for seasonal changes due to RSV and other respiratory virus outbreaks and Rotavirus and other viral causes of AGE, we included two pairs of sine and cosine terms (Fourier terms) in the models to account for seasonality. This approach takes account of pre-lockdown trends and allows estimation of the effect of each level of lockdown and whether there was a change in admissions during the period following the cessation of all lockdowns post-COVID. We built separate models by age (under one year, under five years and between 5 and 13 years) and diagnosis (LRTI, AGE and SAM). Age-specific changes thus do not sum to the total change because the total admissions (and deaths) were analysed as a separate time series. We checked for auto-correlation by calculating the auto-correlation and partial autocorrelation functions. We analysed data using R4.0 (R Foundation for Statistical Computing, Vienna, Austria; appendix).

## RESULTS

During the 60-month study period that extended from 01 April 2018 to 31 March 2023, 45 015 children were admitted across all four specialist hospitals in Durban (eThekwini district). Of these, 20·490 (45·5%) were <1 year of age(infants), 16 549 (36.8%) were children between one and below five years, and 7976(17·7%) were children between five and below 13 years. Across all these age groups, 1237 children died in hospital during the 60 months of the study period, with 733(59·3%) being infants, 346(28%) between one and below five years and 158(12·7%) between five and 13 years. Table 1 compares unadjusted mean monthly admission and mortality numbers and raw case fatality rates during the three assessed periods. While the mean monthly admission appeared marginally lower in the COVID-19 period, there was less of a decrease in mean monthly deaths. The case fatality rates for LRTI, AGE and SAM in the under-five-year group were higher during COVID-19.

**Table 1.** Unadjusted mean monthly admission and mortality numbers during all lockdown levels and post-COVID period.

### Interrupted time series analysis

#### Adjusted admission and mortality monthly numbers by age group

The analysis showed a significant decrease in total admissions for children under 1, 1-to-5-year-olds, and 5-13-year-olds. Level 5 lockdowns saw the most significant mean monthly decreases of 450(95% CI=-657·3–-244·3) p<0·001, 213.2(-349–-76·8) p=0·003, 376·4(- 566·3–-186·4) p<0·0001 in total, under-1-year-old and 1-to-5-year-old admissions respectively. Level-1-lockdowns had the lowest mean monthly decreases. The trend was similar for school-going children (5-13-year olds) to all other age groups. There was also evidence of seasonality on a 6-month scale during the pre-and post-COVID periods in total admissions (p=0·002), under-1-year-olds (p=0·034) and 1-to-5-year olds (p=0·004) which was not evident during the COVID-19 lockdown periods. In the segmented regression model, there was no evidence that excess monthly mortality in SA was associated with changes in admission numbers in any age group. Figures 1(a),1(b),1(c) and 1(d) illustrates these findings.

**Figure 1.**
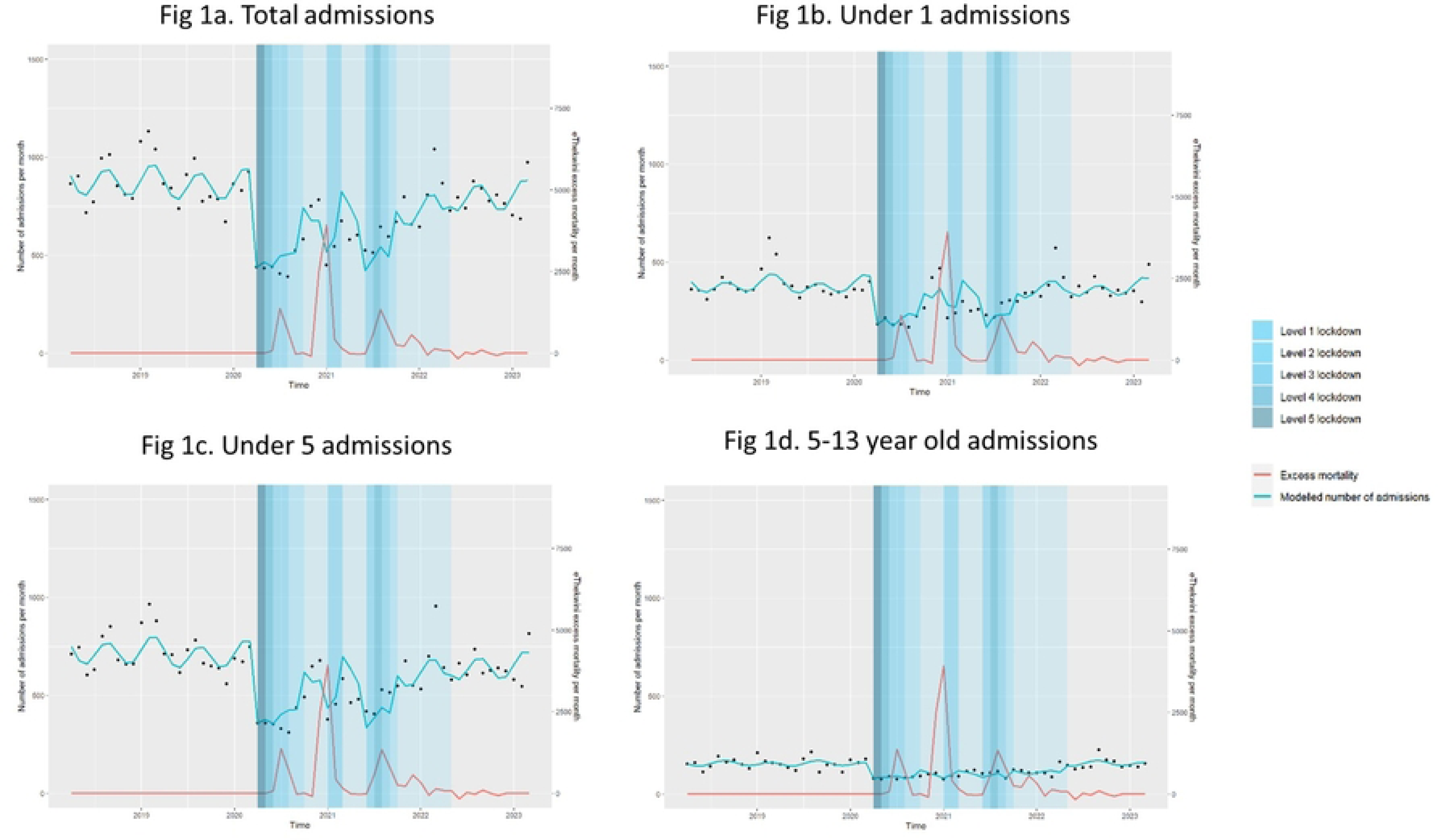
Interrupted time series analysis of admissions by age group, all ages 1(a), under 12 months 1(b), under 60 months 1(c) and between 5 and 13 years old (1d).

In the post-COVID-19 period, total admissions remained slightly lower than during the pre-COVID-19 period (decrease of 68, 95% CI=-134·2–-2·4, p=0·0430). This was mainly due to a decrease in the 1-5-age group (-60·9, 95% CI=-121·5–0·2, p=0.049), with no evidence of a difference in the under-1-age group (+6.5, −50·1–37·0, p=0·765), nor 5-13-year olds (-9·6, - 26·8–7.7, p=0·270).

The segmented regression analysis showed no significant change in monthly mortality in all ages nor specifically in the age categories of under-1-year-olds and 1-to-5-year-olds and 5-13-year age groups during any lockdown levels, nor the post-COVID period. (Table 2, Figure 2(a), 2(b), and 2(c) provide the data and illustrate the trends, respectively)

**Figure 2.**
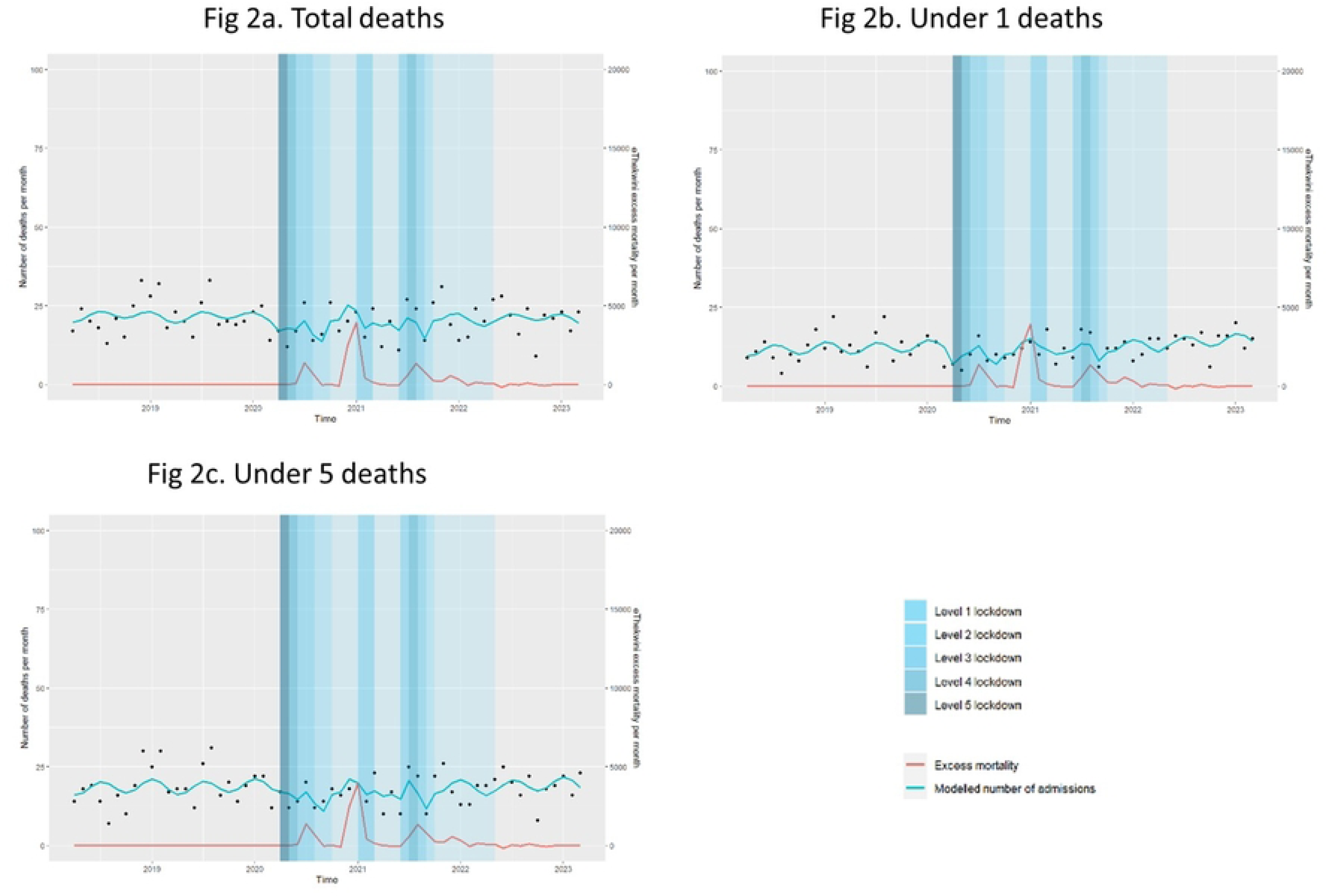
Interrupted time series analysis of mortality rates by age group all ages 2(a), under 12 months2(b) and under 60 months 2(c).

**Table 2.** Changes in mean monthly admissions and mortality in lockdown levels 1 to 5 in all age groups.

#### Admission and mortality rates for children with acute gastroenteritis (AGE), lower respiratory tract infections (LRTI)and severe acute malnutrition (SAM)

Significant decreases in admisssions were seen during most of the lockdown levels in the COVID-19 period in children hospitalised with AGE, LRTI or SAM (Table 3). Level 5 lockdowns saw decreases of 82·8(95%, CI=156·3–-9·3) p=0·028, 132·8(-238·6–- 27·0)p=0·015 and 25·7(95%, −47·4–-3·9), p=0·022 in AGE, LRTI and SAM cases. The terms for seasonality provided evidence of seasonal variation on both a 6-month (p=0·032) and 12-month (p=0·003) scale for AGE admissions, a 6-month scale for LRTI admissions (p=0·004) and a 12-month scale for SAM admissions (p<0·001).

**Table 3.** Changes in adjusted mean monthly admission and case mortality numbers in 1-5-year-old children with Acute Gastroenteritis and Lower Respiratory tract infections during all lockdown levels and post-Covid-period.

Figures 3(a), 3(b) and 3(c) illustrate these changes and loss of the seasonal patterns in AGE and LRTI seen during the COVID-19 period.

**Figure 3.**
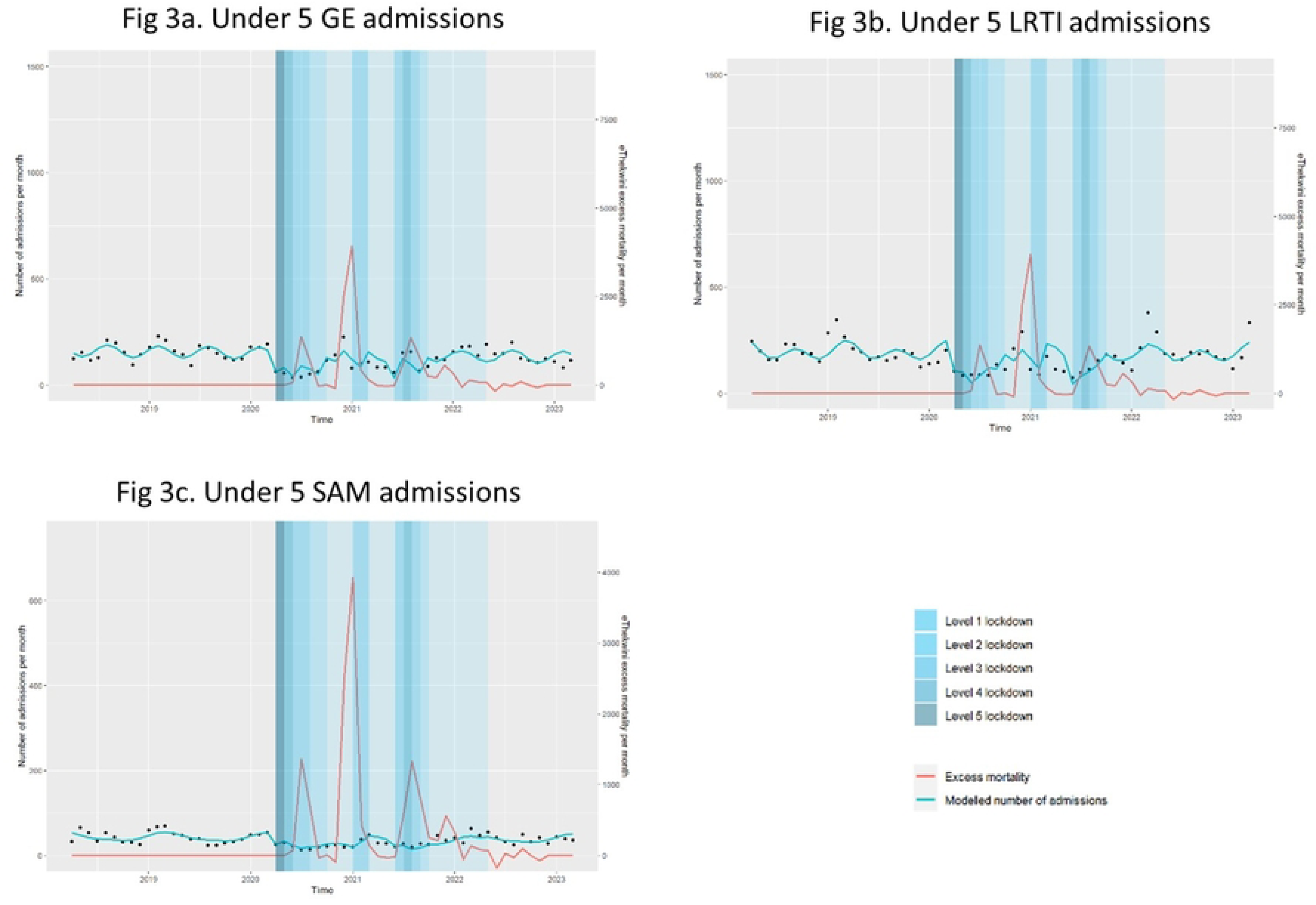
Interrupted time series analysis of admissions by diagnosis in 1-5 year old with acute gastroenteritis 3(a), lower respiratory tract infections 3(b) and severe acute malnutrition 3(c).

In the post-COVID period, there was no evidence that AGE, LRTI, or SAM admissions changed compared to pre-COVID numbers. Figures 3(a), 3(b) and 3(c) illustrate a return to seasonal patterns in the post-COVID period for cases of AGE and LRTI.

When analysing changes in mortality in those hospitalised with either AGE, LRTI or SAM, no significant changes were noted in all the lockdown levels. Table 3 and Figures 4(a), 4(b) and 4(c) illustrates these findings.

**Figure 4.**
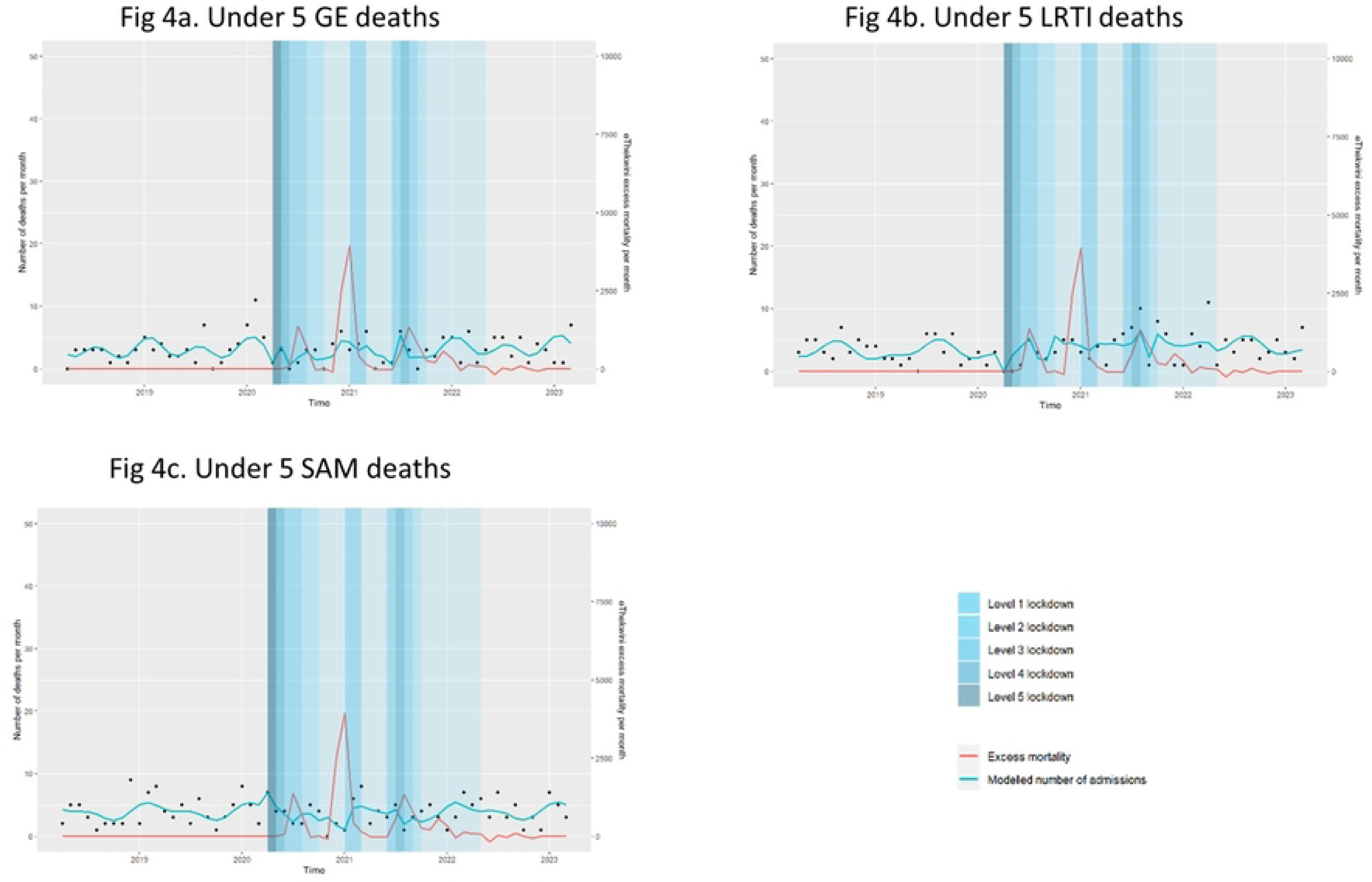
Interrupted time series analysis of mortality rates per admission diagnosis in 1-5 years-olds with acute gastroenteritis 4(a), lower respiratory tract infections 4(b) and severe acute malnutrition 4(c).

## DISCUSSION

Our analysis shows that changes in patterns of admissions and mortality of vulnerable SA children following the COVID-19 pandemic does differ from experiences elsewhere in the world. Despite significant decreases in admissions and changes in seasonal patterns of communicable diseases during the COVID-19 lockdowns, there was neither a concomitant decrease in in-hospital deaths nor was there an anticipated post-pandemic surge in admissions in children from communities with high levels of HIV,TB and poverty.

Several modelling studies and early reviews from LMICs have raised concerns about the impact of the COVID-19 pandemic on vulnerable populations, especially those where fragile healthcare systems exacerbate delayed access to care[10,25]. In our study reflecting sick children requiring hospital admission and drawn from low-income communities, a high population density and existing infectious burden admission numbers did decrease, as was documented in high-income countries, following the promulgation of stringent lockdowns [5,13,15,26]. These decreases in admissions at referral hospitals mirrored decreases in admissions and visits to primary health clinics [4]. Of concern, however, is that the documented decrease in primary care visits and referral hospital admissions could reflect decreased access to healthcare for sick children. Whilst lockdown laws permitted the seeking of healthcare and all facilities remained open through the COVID-19 pandemic, the significant decreases in the admission of sick children, raises the likelihood of worsening of access to health care amongst vulnerable populations. In addition to concerns with decreased access to health care these findings may reflect the influence of a decreased transmission of common childhood communicable diseases, possibly affected by decreased social interactions and mitigating strategies to prevent COVID-19 transmission [9,10]. It has been postulated that increased preventative hygiene habits adopted during the COVID-19 period, like masking, regular hand washing, creche and school closures, and other restrictions impacting person-to-person spread of infections, resulted in modified seasonal patterns of communicable diseases like Rotavirus associated AGE and Respiratory syncytial Virus associated LRTI[6,8,27]. The impact of this possible outcome however has not been fully understood in vulnerable child populations including those with high population densities.

In this study, that reflects children admitted at referral hospitals, including those with complex problems and diagnoses, mortality numbers in all age groups and children with AGE, LRTI and SAM did not decrease during the lockdown period, unlike previously reported [10]. Our finding of a persistence of high mortality despite significant decreases in admissions in the COVID-19 period has been documented elsewhere in poor socio-economic communities [3]. Concerns that this reflects an overall increase in child mortality is not borne out however by any significant increase in excess childhood mortality as seen in age-specific annualised excess death rates (per 1000 population) documented over this period [28].

We postulate that in our large cohort of children hospitalised in public sector referral hospitals there are many children especially those living within high population densities that continued to have exposure to many childhood infections and continued to have a delayed access to care for a multiplicity of reasons. This latter group has been previously documented as experiencing delays in accessing standard healthcare despite the availability of free public health services[29]. Many caregivers here are noted to utilise multiple other sources of care, including allopathic, indigenous and home treatments prior to presenting at public services often with severe complication or in severe distress [29]. It is possible that caregivers in this sub-group would have persisted with late presentation for acute care, similar to pre-pandemic behaviours or delayed their access to hospital care even later. Further exploration is thus required to determine how this vulnerable group were uniquely affected by the challenges posed both by the COVID-19 pandemic and the associated lockdowns.

Our study also documents that the expected surge in malnutrition cases during the lockdown period did not occur, unlike those reported in other studies from developing countries[10,30]. The persistence of high mortality rates in SAM similar to pre-pandemic levels cases despite the decreased admission also suggests that sick children did get to healthcare; however, they could have done so later than was previously the case [28].

The great concern expressed across the world following the decreased utilisation of preventative services and, specifically for vulnerable populations where TB and HIV are endemic, was an expected post-pandemic surge[11,25]. With the disruption of seasonal patterns of viral bronchiolitis, an expected surge in LRTI was also anticipated and documented across countries with severe lockdown [31,32]. In this study, which analysed data over a longer post-pandemic period than most other studies, we do not show this anticipated surge in admissions in under-1-year and 1-to-5-year-old children as well as in cases of AGE, LRTI and SAM. The trend identified in the post-pandemic period may reflect a gradual increase in admissions back to pre-COVID levels. Our postulates is that these vulnerable children living high population densities with cramped living, were exposed to common childhood infections unlike these children living in low populations densities the lack of a post-pandemic surge seen with most of the children in this cohort could be explained by this possibility.

Strengths of our study include the large cohort of children hospitalised specifically for medical diagnoses in public sector referral hospitals. We reflected on acutely sick and vulnerable children susceptible to communicable diseases. Our use of long-term routine data considers pre-COVID, all the COVID lockdown levels, and substantial post-COVID periods, whilst most studies have largely focused on the COVID-19 period. However we were not able to verify definitive microbiological, virologic and formal HIV and TB results; instead, we relied on retrospective diagnoses provided by source documents and by paediatricians on site. Further studies specifically targeting these populations with verifiable microbiological testing may be required to unpack children’s behaviours under differing contexts. This study may help determine the epidemiological patterns of vulnerable children when faced with communicable disease outbreaks in greater detail. We did not focus on neonatal or non-medical or children admitted to referral intensive care units (ICU) requiring prolonged ventilation. We could not assess the definitive socio-economic status and inferred this based on previous usage patterns in public sector hospitals. The retrospective data reflects in-hospital mortality specifically and does not include community-based death data.

In conclusion, our findings suggest that, in one of the regions most affected by HIV, Tuberculosis and malnutrition whilst admissions of acutely sick children decreased similar to other countries with better health resources, a decrease in in-hospital mortality and anticipated post-pandemic surges in admission was not seen as compared with these countries. This study provides evidence that children, in vulnerable communities with high population densities, HIV and TB infection rates, behaved differently to communities where these conditions were not as common. These findings suggest that mitigating strategies to reduce infectious disease outbreaks possibly affected transmission dynamics of common communicable childhood diseases, differently in communities and this requires further exploration and study. Further studies in vulnerable populations are needed to identify persisting challenges in healthcare provision, infection transmission dynamics and the impact of promulgation of uniform pandemic control measures on child health outcomes.

## DECLARATIONS

## Acknowledgements

The authors would like to express their gratitude to Mrs Leora Sewnarain’s assistance with formatting and language review.

## Availability of data and materials

The data used for this analysis cannot be shared publicly because of legal and ethical requirements regarding using routinely collected clinical data in South Africa. Researchers may request access to the data from the eThekwini Municipality Health Unit and Kwa-Zulu Natal Department of Health (contact details obtainable upon request to the corresponding author).

## Authors’ contributions

KLN, JD and MA conceptualised the study. KLN, KC, ML and YN oversaw data collection. KLN, KC, ML and YN oversaw the curation of the data. JD, CB and KLN analysed the data. KLN drafted the manuscript. JD, CB and KLN verified the underlying data. All authors critically reviewed and edited the manuscript and consented to final publication. KLN and JD had full access to all the data, and all authors had final responsibility for the decision to submit for publication.

## Disclaimer

The views and opinions expressed in this article are those of the authors and do not necessarily reflect the official policy or position of any affiliated agency of the authors.

## Ethical consideration

Adherence to ethical guidelines was ensured throughout the research process. The study was approved by the University of KwaZulu-Natal Biomedical Research Ethics Committee (BREC/00002981/2021), the KwaZulu-Natal Department of Health’s Provincial Health Research Ethics Committee, eThekwini District Health Department and the Child Health Identification Programme (National committee) with a waiver for informed consent for analysis of anonymised, routinely collected data.

## Consent for publication

Not applicable.

